# Diagnostic value in colorectal tumorous lesions of *SDC2/TFPI2* methylation based on bowel subsite difference

**DOI:** 10.1101/2021.02.22.21252188

**Authors:** Lianglu Zhang, Lanlan Dong, Changming Lu, Wenxian Huang, Cuiping Yang, Qian Wang, Qian Wang, Ruixue Lei, Rui Sun, Kangkang Wan, Tingting Li, Fan Sun, Tian Gan, Jun Lin, Lei Yin

## Abstract

**Background:** *SDC2* methylation is a potential biomarker for colorectal cancer detection with specificity over 90%, but its sensitivity is usually less than 90%. The study aims to improve the sensitivity and specificity by adding *TFPI2* methylation as a complement.

**Methods:** *TFPI2* was identified using colorectal cancer samples from TCGA database with *SDC2* methylation level lower than 0.2. *SDC2*/*TFPI2* methylation specific PCR was performed using tissue samples (184 cancer and 54 healthy control) and stool samples (289 cancer, 190 adenoma and 217 healthy control). Detection sensitivity was calculated among tumors from different biopsy locations.

**Results:** 88% of 50 TCGA specimens of colorectal cancer with *SDC2* methylation level lower than 0.2 showed *TFPI2* methylation level higher than 0.2. *SDC2*/*TFPI2* combined detection in stool specimens showed AUC value of 0.98 with the specificity of 96.40% and sensitivity of 96.60% for cancer vs control, AUC value of 0.87 with the specificity of 95.70% and sensitivity of 80.00% for adenoma vs control. The sensitivities were much higher than those of *SDC2*, and the improvement was most significant in lesions from left colon and rectum.

**Conclusions:** This research indicated that the addition of *TFPI2* can reduce the miss detection rate of colorectal cancer and adenomas while maintaining high specificity, probably by finding neoplasms in left colon.The method is non-invasive and has good compliance, also avoiding the pain of bowel preparation and risk of cross infection during endoscopy, so it will provide an easy and precise tool for colorectal cancer and its precancerous lesions screening.

## 1 Introduction

Colorectal cancer (CRC) affects millions of people around the world. It’s unique because of slow progress, making it preventable and often curable [1,2]. The five-year survival rate can be as high as 90% or less than 10%, depending on the stage of diagnosis [3]. Sporadic CRC mainly develops in a normal - adenomas - carcinoma sequence [4] and early detection of CRC can significantly decrease mortality [5]. Colonoscopy is the gold standard for CRC diagnosis at present [6], but due to the invasive and complex intestinal preparation process, its compliance in the average risk population is low [7]. Stool occult blood test (FOBT) and stool immunochemistry test (FIT) are two noninvasive methods, but their sensitivity is relatively insufficient, especially for stage I CRC and advanced adenomas [8]. The occurrence of CRC is related to genomic and epigenetic changes, such as gene mutation, chromosome instability, microsatellite instability and CpG island aberrant methylation [9]. Among them, CpG island methylation is the most common change during tumorigenesis [10].

Aberrant DNA methylation can occur at the very early stage of cancer [11], therefore, by distinguishing the methylation patterns difference between malignant and normal samples, very useful biomarkers can be found. So far, several DNA methylation biomarkers for CRC have been identified, including *SDC2, NDRG4, BMP3, VIM, SFRP2*, and *SEPT9* [12-17], but the sensitivity of a single marker is usually lower than 90% [18].The first stool-based CRC detection product “Cologuard”, targeting the hemoglobin, *KRAS* mutation and two methylated genes (NDRG4 and BMP3), has a sensitivity of 92% and specificity of 87% for CRC [19], however, multiple targets tests may be costly and difficult to implement.

*SDC2* has been identified as a potential biomarker for CRC using a genome-wide CpG microarray approach [16]. Aberrant methylation in *SDC2* CpG islands has been found in different types of biological specimens like tissue, blood and stool [16, 20]. A study based on Chinese stool samples showed that the sensitivity and specificity of *SDC2* methylation for CRC was 81.1% and 93.3%, respectively [21]. To improve the sensitivity, Korean researchers adopted an LTE-q methylation-specific PCR (MSP) method to enrich *SDC2* by two-round PCR, and the overall sensitivity reached 90% for CRC [22, 23]. However, it might inevitably increase experimental complexity and time cost.

In this study, we chose an alternative path to improve the sensitivity of *SDC2* in MSP assays. Through genome-wide screening, we found that *TFPI2* gene was highly complementary to *SDC2* for CRC and adenomas detection. The combined detection of *TFPI2* and *SDC2* showed both high specificity and sensitivity, especially for cancer and adenomas in left colon both for tissue and stool specimens. Here we present the results of *TFPI2* identification, and the evaluation of MSP systems in tissue and stool of patients with colorectal lesions at different bowel sites.

## 2 Methods

### 2.1 Patients and sample collection

This study was approved by the Medical Ethics Committee, Zhongnan Hospital of Wuhan University (ethical approval No.2019099). Tissue specimens of 184 CRC patients and 54 healthy controls (Supplemental Table 1) were collected from Zhongnan Hospital of Wuhan University, and stool specimens of 289 CRC patients, 190 adenomas patients, and 217 healthy controls (Supplemental Table 2) were from Zhongnan hospital and Renmin hospital of Wuhan University, Ruijin hospital of Shanghai Jiaotong University, Wuhan eighth hospital of Hubei University of Chinese Medicine, the fourth affiliated hospital of Henan University of Science and Technology, Wuhan fourth hospital of Huazhong University of Science and Technology. Written informed consent was obtained from all participants.

**Table 1.**
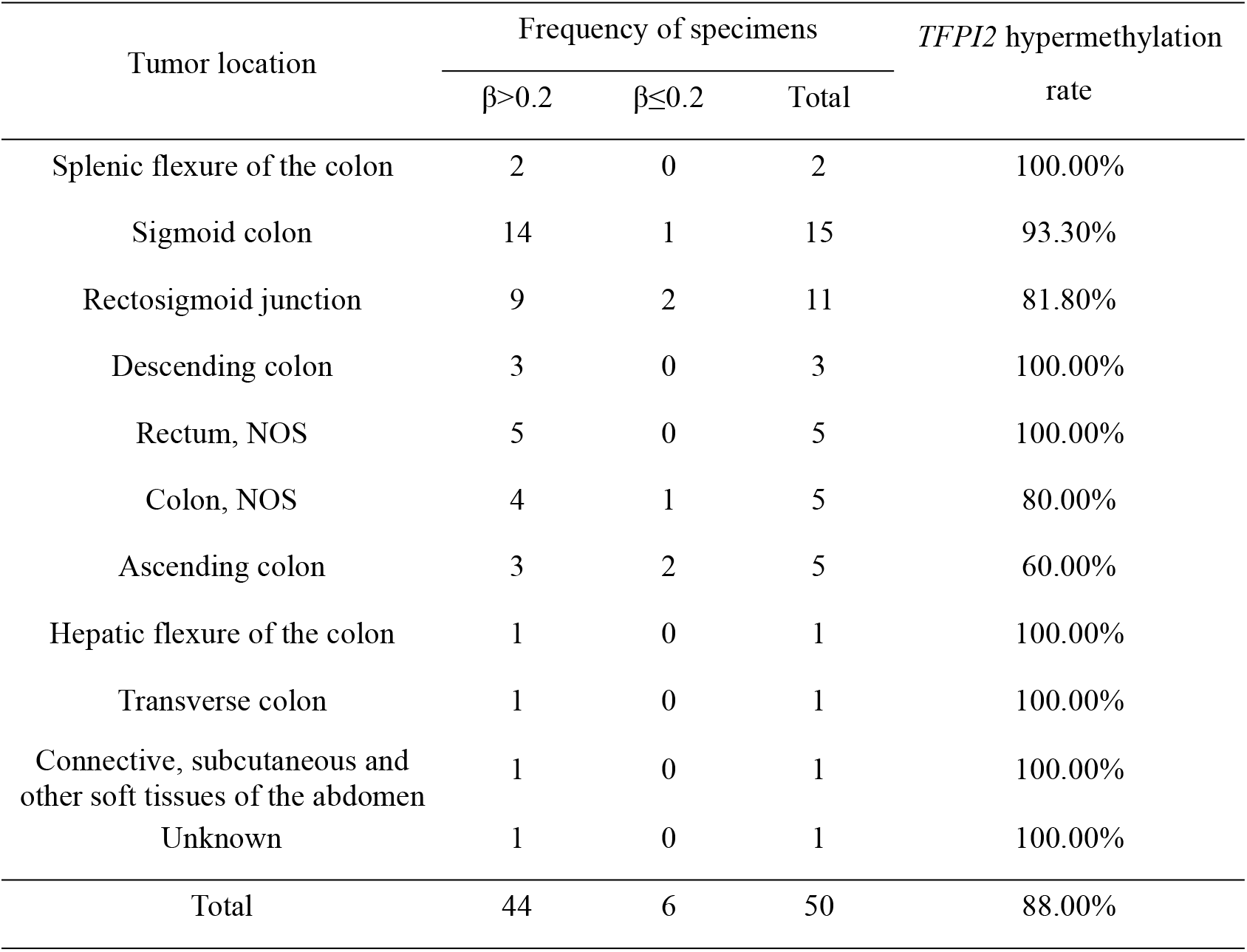
Detection rate of *TFPI2* in colorectal cancer tissues with *SDC2* β value lower than 0.2

**Table 2.** Methylation-specific PCR results in stool specimens with Ct-value and ΔCt as an indicator*

| Marker | Indicator | Specificity | Sensitivity | AUC | AUC 95% CI | Group |
| --- | --- | --- | --- | --- | --- | --- |
| SDC2 | Ct | 99.30% | 44.20% | 0.64 | 0.57 to 0.71 | Adenoma vs.<br>normal |
| TFPI2 | Ct | 100.00% | 67.40% | 0.85 | 0.81 to 0.89 |  |
| SDC2/TFPI2 | Ct | 95.70% | 80.00% | 0.87 | 0.81 to 0.92 |  |
| SDC2 | Ct | 100.00% | 78.60% | 0.88 | 0.85 to 0.92 | Cancer vs.<br>normal |
| TFPI2 | Ct | 100.00% | 88.80% | 0.95 | 0.93 to 0.97 |  |
| SDC2/TFPI2 | Ct | 96.40% | 96.60% | 0.98 | 0.96 to 0.99 |  |
| SDC2 | Ct | 54.70% | 81.60% | 0.69 | 0.63 to 0.75 | Cancer vs.<br>adenoma |
| TFPI2 | Ct | 32.60% | 90.80% | 0.63 | 0.59 to 0.70 |  |
| SDC2/TFPI2 | Ct | 64.20% | 73.80% | 0.72 | 0.67 to 0.78 |  |
| SDC2 | $\Delta$ Ct | 73.20% | 45.30% | 0.51 | 0.42 to 0.59 | Adenoma vs.<br>normal |
| TFPI2 | $\Delta$ Ct | 89.90% | 71.60% | 0.78 | 0.70 to 0.84 | |
| SDC2/TFPI2 | $\Delta$ Ct | 92.80% | 69.50% | 0.78 | 0.72 to 0.85 | |
| SDC2 | $\Delta$ Ct | 86.20% | 56.80% | 0.72 | 0.66 to 0.77 | Cancer vs.<br>normal |
| TFPI2 | $\Delta$ Ct | 89.90% | 78.60% | 0.86 | 0.82 to 0.90 | |
| SDC2/TFPI2 | $\Delta$ Ct | 90.60% | 82.00% | 0.87 | 0.84 to 0.90 | |
| SDC2 | $\Delta$ Ct | 75.80% | 53.40% | 0.67 | 0.61 to 0.72 | Cancer vs.<br>adenoma |
| TFPI2 | $\Delta$ Ct | 71.60% | 55.30% | 0.63 | 0.56 to 0.69 | |
| SDC2/TFPI2 | $\Delta$ Ct | 78.90% | 53.40% | 0.67 | 0.62 to 0.74 | |

\*The cutoff values were based on the maximized Youden index.

All MSP tests were done by investigators blinded to patients’ colonoscopy and pathology information, MSP results were analyzed independently by other researchers.

### 2.2 Discovery of biomarker complementary to *SDC2*

391 CRC specimens from the cancer genome atlas (TCGA) were sorted according to the methylation level (mean β value of the probes within CpG island) of *SDC2* to identify hypomethylated specimens (methylation level≤0.2). Hypermethylated genes (methylation level>0.2) were identified among hypomethylated specimens through whole-genome analysis. Receiver operating characteristic (ROC) analysis were performed using methylation level of each sample to evaluate the diagnostic complementarity of hypermethylation genes and *SDC2* in different colorectal sites.

### 2.3 Specimen processing and DNA extraction

Genomic DNA of tissue specimens was isolated using QIAamp DNA FFPE Tissue Kit (Qiagen, Hilden, Germany) according to the manufacturer’s instruction.

8 g stool specimen and kept with 32 mL preservation buffer (200 mmol/L Tris·HCl, 300 mmol/L EDTA·2Na, 150 mmol/L NaCl, pH 8.0) for DNA isolation, the biotin-labeled capture probes were designed for *SDC2, TFPI2*, and reference gene *ACTB*, sequences were shown in Supplemental Table 3. After centrifugation, DNA was denaturated under 90°C for 15 min. The single-strand DNA and the capture probes were then incubated with streptavidin magnetic beads at room temperature for 1 h. After washing twice, the target DNA was eluted with a 50 uL TE buffer.

**Table 3.** Detection rate of *TFPI2* in stool specimens with *SDC2* Ct value higher than 38

| Type of samples | Tumor location | Frequency of specimens |  |  | *TFPI2* detection rate |
| --- | --- | --- | --- | --- | --- |
|  |  | Positive | Negative | Total |  |
| Carcinoma | Left colon | 5 | 3 | 8 | 62.50% |
|  | Right colon | 1 | 3 | 4 | 25.00% |
|  | Sigmoid colon | 8 | 5 | 13 | 61.50% |
|  | Rectal | 7 | 6 | 13 | 53.80% |
|  | Total | 21 | 17 | 38 | 55.30% |
| Adenoma | Left colon | 5 | 12 | 17 | 29.40% |
|  | Right colon | 1 | 16 | 17 | 5.90% |
|  | Sigmoid colon | 8 | 15 | 23 | 34.80% |
|  | Rectal | 7 | 19 | 26 | 26.90% |
|  | Total | 21 | 62 | 83 | 25.30% |
| Normal | Total | 6 | 202 | 208 | 2.89% |

### 2.4 Bisulfite Conversion

DNA was converted using EZ Methylation Kit (Zymo Research. LA, America) according to the manufacture’s instruction. The eluted DNA was either used immediately or stored at -20°C for further use.

### 2.5 MSP

Sequences of primers and probes were shown in Supplemental Table 3. PCR solution was prepared in a volume of 25 uL with High-Affinity Hotstart Taq Polymerase (TIANGEN, Beijing, China). 5uL template DNA was added, non-template control, methylated and non-methylated controls were tested together in every plate. PCR was performed on ABI 7500 instrument under the following cycling conditions: 95 °C for 10 min, followed by 45 cycles of 95 °C for 15 s and 60 °C for 30 s.

MSP results were utilized in ROC analysis. The optimal cutoff was determined by maximizing Youden’s index. The area under the ROC curve (AUC) value, sensitivity and specificity were estimated.

When calculating the detection effect of marker/s of cancer and adenoma in different colorectal sites, the cutoff value was set at Ct=38 for *SDC2* or *TFPI2* and Ct=36 for *ACTB*. If the Ct value of *ACTB* >36, the reaction was invalid. The specimen was methylation positive if the Ct value of *SDC2* or/and *TFPI2* ≤ 38. Sensitivity and specificity values were calculated as:

Sensitivity = true positives / total case number ×100%

Specificity = true negatives / total control number ×100%

### 2.6 Statistical Methods

All the bioinformatic and statistical analyses were performed with R version 3.6.1. To determine the statistical significance of the difference in methylation level between case and control group, Wilcoxon signed-rank test was used for pair wised data and Mann-Whitney U test for group wised data. Sensitivities between different colorectal locations was tested by the Fisher exact test.

## 3 Results

### 3.1 High frequency of hypermethylated *TFPI2* is present in *SDC2* hypomethylated CRC specimens

The identification of *TFPI2* was achieved by analyzing the 450K chip data of CRC and normal samples in the TCGA database, the TCGA dataset included 411 case and 45 normal specimens, 391 CRC specimens and 45 normal specimens were retained for marker discovery after removing those of incomplete clinical information and tumor metastasis (Supplemental Table 4). 391 CRC specimens from TCGA were sorted according to the methylation level (mean β value of the probes) of the *SDC2* CpG island. 50 CRC specimens had a methylation level lower than 0.2 (Supplemental Table 5). Genes with β value greater than 0.2 in these *SDC2* hypomethylated specimens were retrieved throughout the whole genome, and genes complementary to *SDC2* in different colorectal sites were selected, *TFPI2* showed the best complementarities to *SDC2* in various colorectal sites (Table 1). 44 out of 50 specimens showed *TFPI2* β value higher than 0.2, accounting for 88.0% (Table 1).

### *3*.*2 TFPI2* and *SDC2* are heavily hypermethylated in case rather than control specimens in TCGA, GSE48684 and GSE79740 datasets

In order to further evaluate the two markers of *SDC2* and *TFPI2*, we compared the methylation levels of CRC, adenomas and normal samples in the three datasets of TGGA, GSE48684 and GSE79740. GSE48684 and GSE79740 were downloaded from Gene Expression Omnibus (GEO) database [24,25]. GSE48684 contained 106 case and 41 control specimens and GSE79740 contains 44 case and 10 control specimens, they were both used for markers validation (Supplemental Table 6). The results was given in Fig 1.

**Fig 1.**
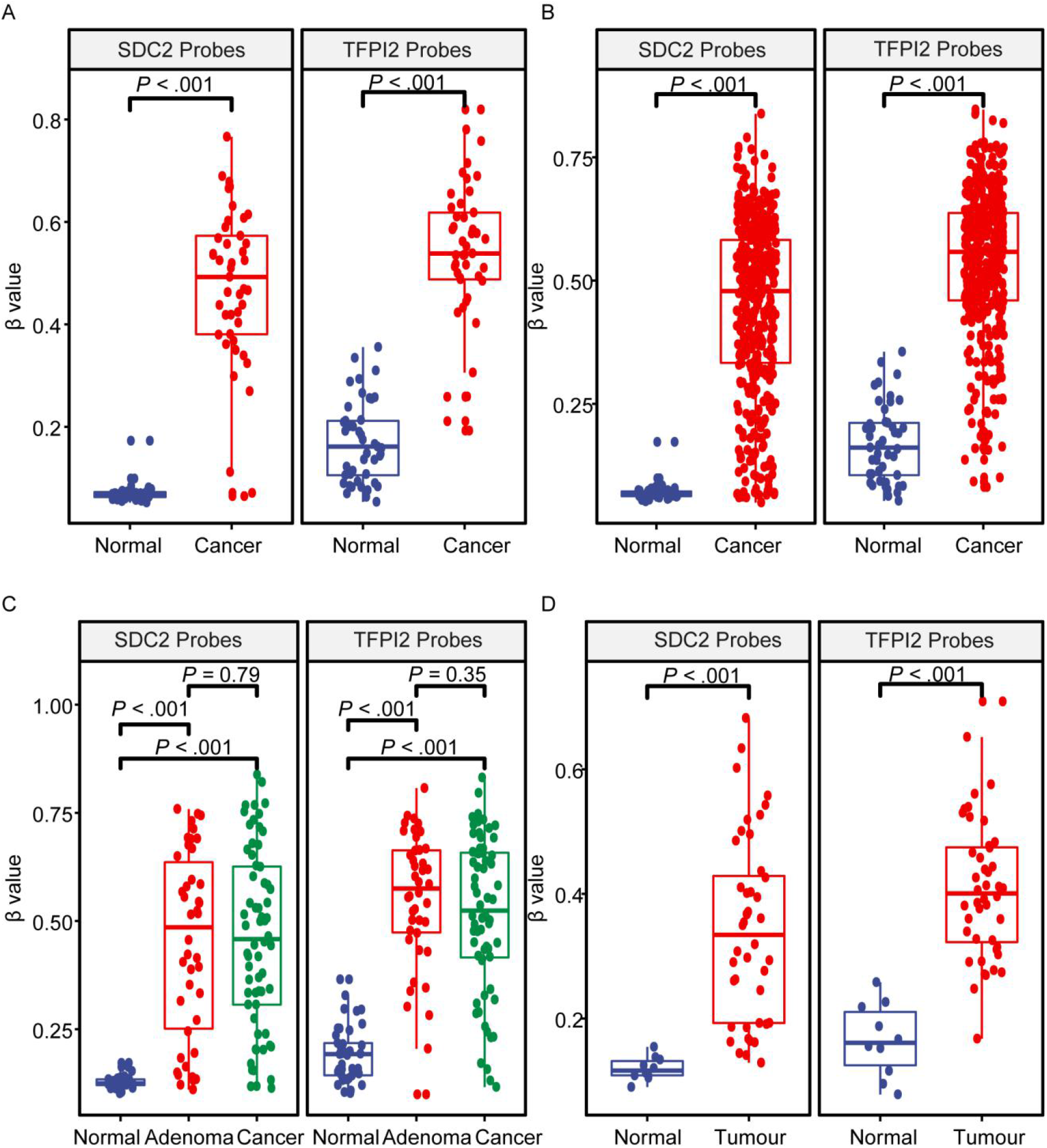
DNA methylation level (β value) of *SDC2* and *TFPI2* in CRC, adenomas, and normal tissues. Fig 1A illustrates the distribution of methylation levels in 45 CRC vs. their paired normal tissue specimens from TCGA. Fig 1B is 391 CRC vs. 45 normal tissue specimens from TCGA. Fig 1C is based on the data set of GSE48684, with 106 CRC and 41 normal tissue specimens. Fig 1D is based on GSE79740, with 44 CRC and 10 normal tissue specimens. Wilcoxon signed-rank test was used for Fig 1A, and Mann-Whitney U test for Fig 1(B-D).

The average methylation level of *SDC2* and *TFPI2* in 45 CRC tissue from TCGA was 0.492 and 0.538, while that in their paired adjacent tissue was 0.067 and 0.161 (Fig 1A), respectively. The difference of either *SDC2* or *TFPI2* methylation level between the 45 paired CRC and normal tissues was highly significant (p<0.001). When a group comparison in methylation level was made between 391 CRC samples and 45 normal samples (Fig 1B), CRC tissue in group was significantly higher than normal tissues, either for *SDC2* (0.479 ± 0.178 v.s.0.067 ± 0.018) or for *TFPI2* (0.558 ± 0.149 v.s.0.161 ± 0.078).

A similar tendency was also observed in the datasets GSE48684 and GSE79740 (Fig 1C&D), the difference was significant between CRC/adenomas and normal groups in this two datasets, furthermore, no significant difference was detected between CRC and adenomas samples (p =0.79 for *SDC2* and p =0.35 for *TFPI2*) (Fig 1C).

### 3.3 *SDC2*/*TFPI2* combined detection has superior discriminatory power of CRC normal Specimens in TCGA dataset

To determine the diagnostic value of *SDC2*/*TFPI2*, ROC curves were analyzed using 391 CRC and 45 normal specimens in TCGA with β = 0.2 as the cutoff, *SDC2*/*TFPI2* combined detection showed higher AUC value (0.99) than *SDC2* (0.95) and *TFPI2* (0.97) (Fig 2). The sensitivity (98.2%) of *SDC2*/*TFPI2* combined detection was also significantly higher than *SDC2* (87.2%) and *TFPI2* (96.4%).

**Fig 2.**
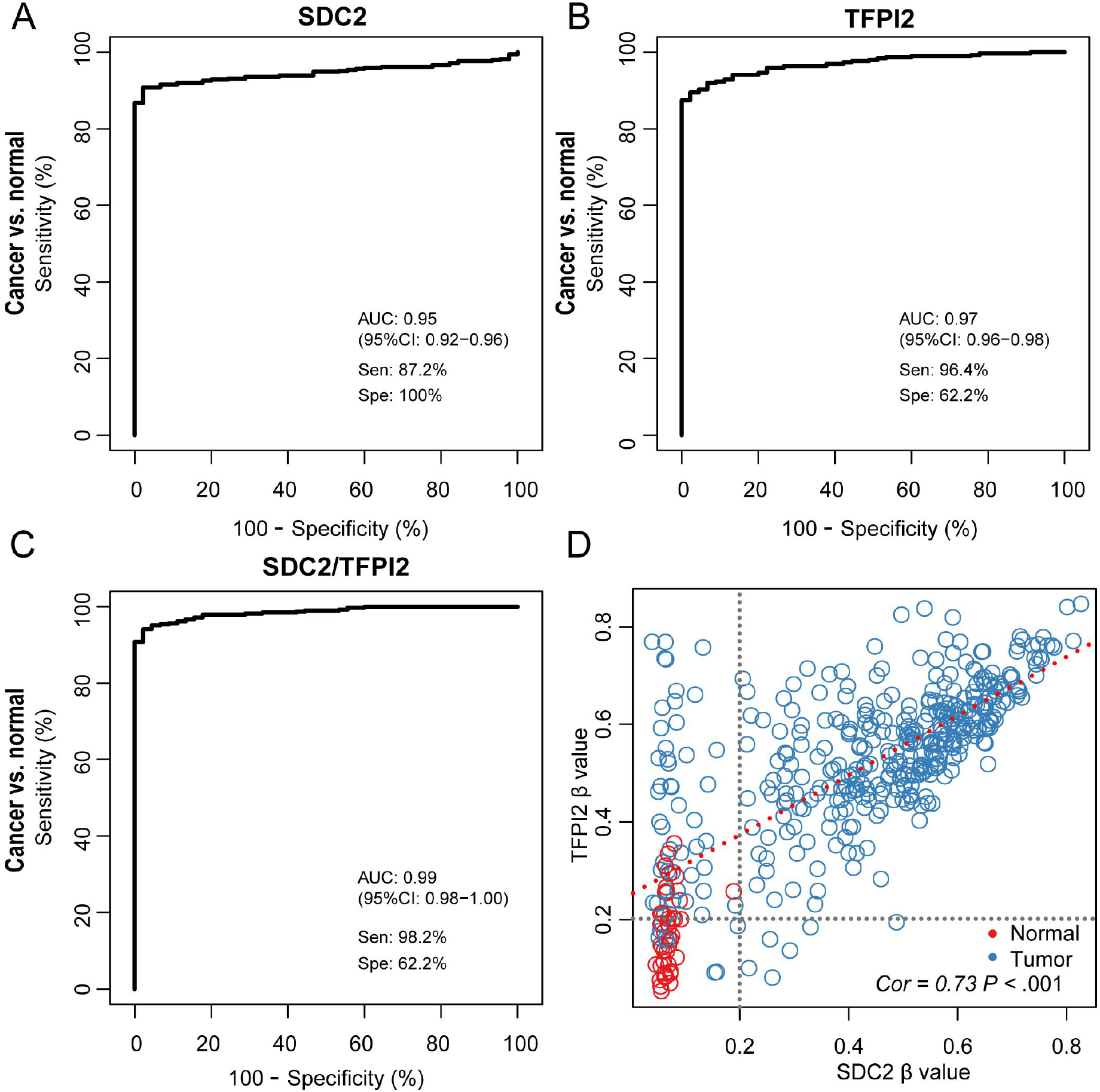
Comparison in diagnostic performance between *SDC2, TFPI2*, and *SDC2*/*TFPI2* using 391 CRC and 45 normal samples from TCGA. Fig 2A is ROC analysis for *SDC2*, Fig 2B for *TFPI2*, and Fig 2C for *SDC2*/*TFPI2* in combination. Fig 2D is a scatter plot of specimens with *SDC2* methylation level as abscissa and *TFPI2* methylation level as ordinate. Cor means the coefficient of correlation between *SDC2* and *TFPI2*. The cutoff value is 0.2 for both *SDC2* and *TFPI2* as indicated by the black dot lines.

With β = 0.2 as the cutoff value of methylation level of *SDC2* and *TFPI2*, the 391 specimens can be divided into four groups (Fig 2D): High *SDC2* High *TFPI2* (High methylation level of *SDC2* and *TFPI2*), High *SDC2* Low *TFPI2* (High methylation level of *SDC2* and low methylation level of *TFPI2*), Low *SDC2* High *TFPI2* (Low methylation level of *SDC2* and high methylation level of *TFPI2*) and Low *SDC2* Low *TFPI2* (Low methylation level of *SDC2* and low methylation level of *TFPI2*). Most tumor specimens (375/391) were allocated in High *SDC2* High *TFPI2* group, High *SDC2* Low *TFPI2* group and Low *SDC2* High *TFPI2* group, while normal specimens were mainly distributed in Low *SDC2* Low *TFPI2* group (44/60). Specimens in High *SDC2* High *TFPI2* group and High *SDC2* Low *TFPI2* group are all tumor specimens without normal tissue specimens involved. All false-positive and false-negative specimens were found in Low *SDC2* Low *TFPI2* and Low *SDC2* High *TFPI2* groups with hypomethylated *SDC2*. Low *SDC2* Low *TFPI2* group were also mixed with a small number of false-negative tumor specimens.

### 3.4 MSP can efficiently differentiate CRC, adenomas and normal specimens based on Ct value

MSP assays were developed and evaluated with 238 tissue (184 CRC and 54 healthy control) and 696 stool specimens (289 CRC, 190 adenomas and 217 healthy control). The methylation level of the target genes was monitored by Ct (Fig 3A&C) and ΔCt values (Fig 3B&D). The PCR reaction was considered as effective if the Ct value of *ACTB* was lower than 36. The endogenous reference gene *ACTB* showed a significant difference in Ct value, and the median Ct values were normal > adenomas > cancer (Fig 3A&C). In order to remedy this difference, all Ct values were normalized into ΔCt values (Ct_*target*_ -Ct_*ACTB*_) for case-control comparison (Fig 3B&D).

**Fig 3.**
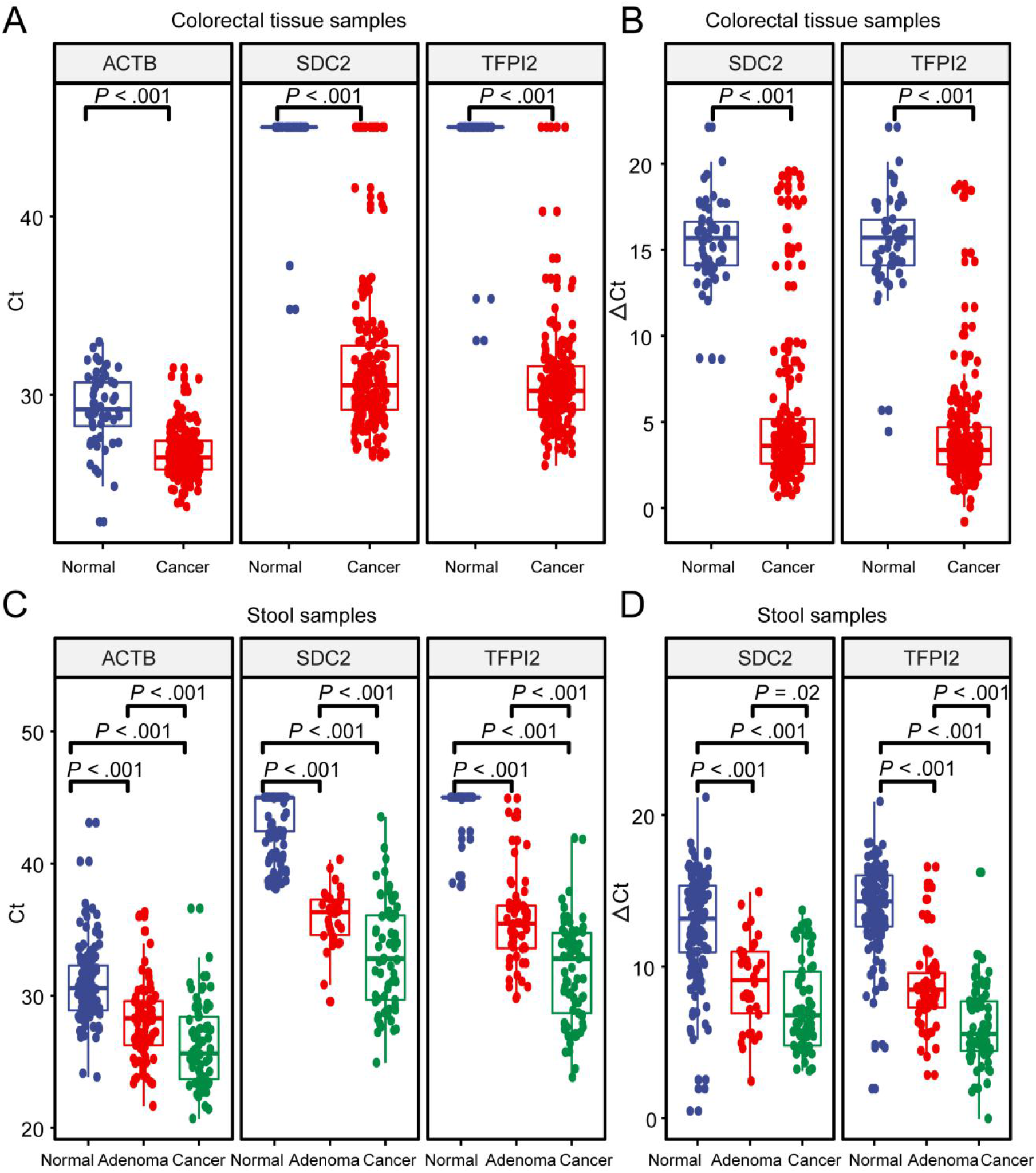
Distribution of Ct and ΔCt values generated by MSP. ΔCt represents the difference between Ct_*Target*_-Ct_*ACTB*_. Fig 3(A&B) are built on colorectal tissue specimens. Fig 3 (C&D) are built on stool specimens. Statistically significant differences were determined using the Mann-Whitney U test.

In tissue specimen testing, Δ Ct value showed normal > adenomas > cancer either for *SDC2* or *TFPI2* (Fig 3B). A highly significant difference was observed between cancer and normal samples (p < 0.001) samples. In stool specimen testing, the Δ Ct values showed the same trend (Fig 3D). A highly significant difference was found between adenomas and normal samples (p < 0.001), and between cancer and normal samples (p < 0.001), the difference between cancer and adenomas was also significant (p =0.02 for *SDC2* and p < 0.001 for *TFPI2*.

### 3.5 *TFPI2* can significantly improve the diagnostic performance of *SDC2* in stool specimens

The diagnostic performance of *SDC2, TFPI2*, and *SDC2*/*TFPI2* in stool samples was evaluated by ROC curve analysis with colonoscopy as the gold standard and MSP result as the evaluation index (Table 2 and Figure 4).

**Fig 4.**
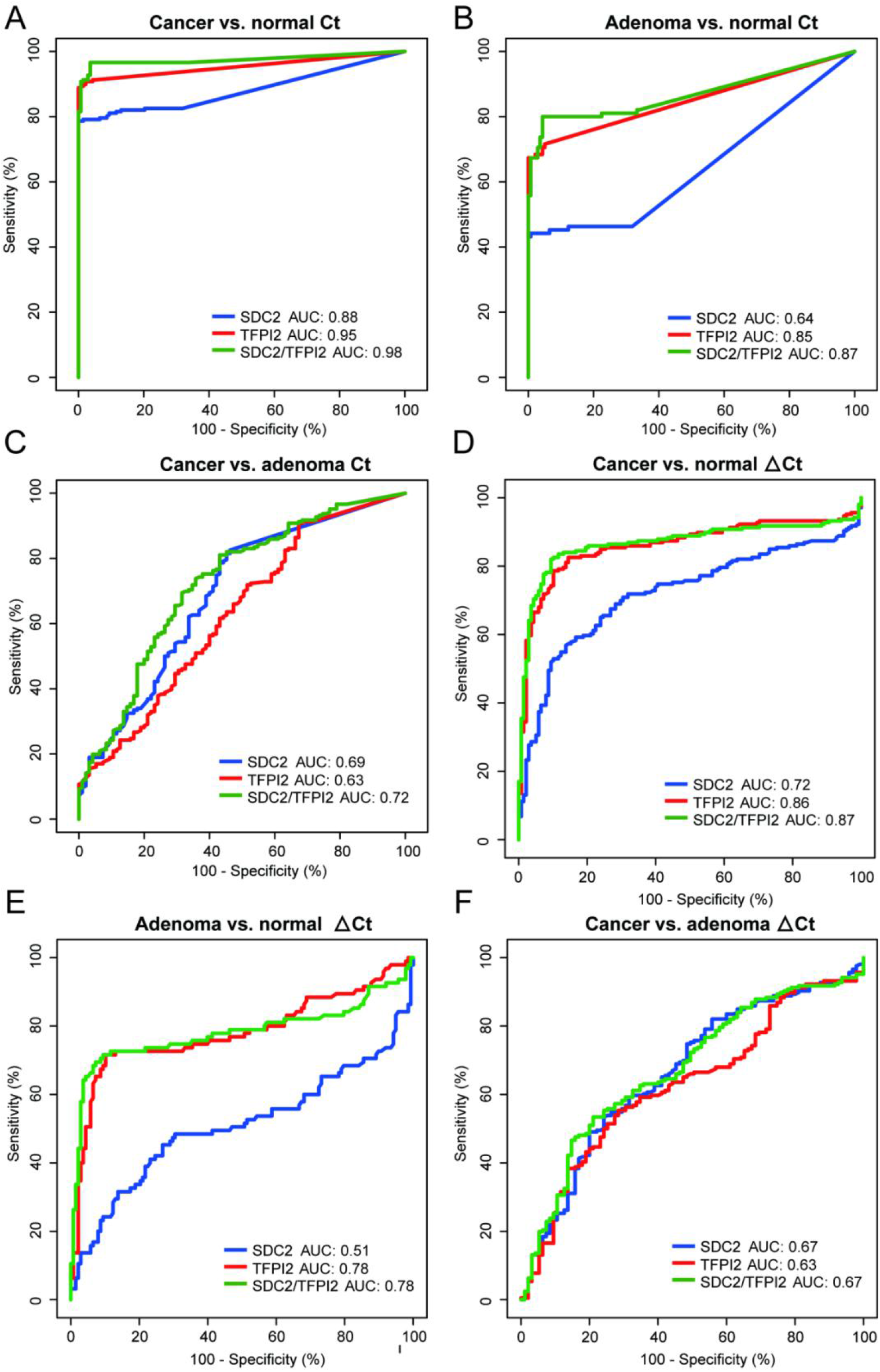
Diagnostic performances of methylation-specific PCR targeting *SDC2, TFPI2*, and *SDC2*/*TFPI2* in stool specimens. Fig 4(A-C) were ROCs based on Ct values, Fig 4(D-F) are based on ΔCt values.

For discrimination between cancer and normal samples based on Ct values, the AUC value of *SDC2*/*TFPI2* combined detection was 0.98 (95% CI: 0.96-0.99) with the specificity of 96.40% and sensitivity of 96.60%, while the AUC value of *SDC2* detection was 0.88 (95% CI: 0.85-0.92) with the specificity of 100% and sensitivity of 78.60%. For discrimination between adenomas and normal specimens, the AUC value of *SDC2*/*TFPI2* combined detection was 0.87 (95% CI: 0.81-0.92) with the specificity of 95.70% and sensitivity of 80.00%, while the AUC value of *SDC2* detection was 0.64 (95% CI: 0.57-0.71) with the specificity of 99.30% and sensitivity of 44.20%. *SDC2*/*TFPI2* combined detection showed better diagnostic performance in distinguishing any two of the three types of specimens.

When Δ Ct values served as methylation level index, *SDC2*/*TFPI2* combined detection showed similar results with Ct-based analysis. However, Δ Ct-based diagnostic performance was significantly lower than the Ct based diagnosis.

### 3.6 *TFPI2* can improve the detection sensitivity of *SDC2* through Finding Cancer in Left Colon, Sigmoid Colon and Rectum

Fig 5 shows the sensitivity comparison of different detection methods (targeting *SDC2, TFPI2* alone and *SDC2*/*TFPI2* jointly) in different biopsy sites. Fig 5A was based on the TCGA dataset with 391 CRC specimens. Sensitivity of *SDC2*/*TFPI2* combined detection was 98.2%, while that of *SDC2* alone was 87.2%, the increase was significant (p ≤ 0.01). Sensitivity difference varied with differen locations, highly significant improvement was found in rectum (n=46, *SDC2*/*TFPI2*=100.0%, *SDC2*=87.0%, p ≤ 0.01), sigmoid colon (n=88, *SDC2*/*TFPI2*=95.5%, *SDC2*=76.1%, p ≤ 0.01) and rectosigmoid junction (n=46, *SDC2*/*TFPI2*=93.5%, *SDC2*=76.1%, p ≤ 0.01). Detection sensitivity was also increased for transverse colon (n=25, *SDC2*/*TFPI2*=100.0%, *SDC2*=88.0%), descending colon (n=14, *SDC2*/*TFPI2*=100.0%, *SDC2*=85.7%) and ascending colon (n=55, *SDC2*/*TFPI2*=98.2%, *SDC2*=94.6%) cancer though they were not statistically significant.

**Fig 5.**
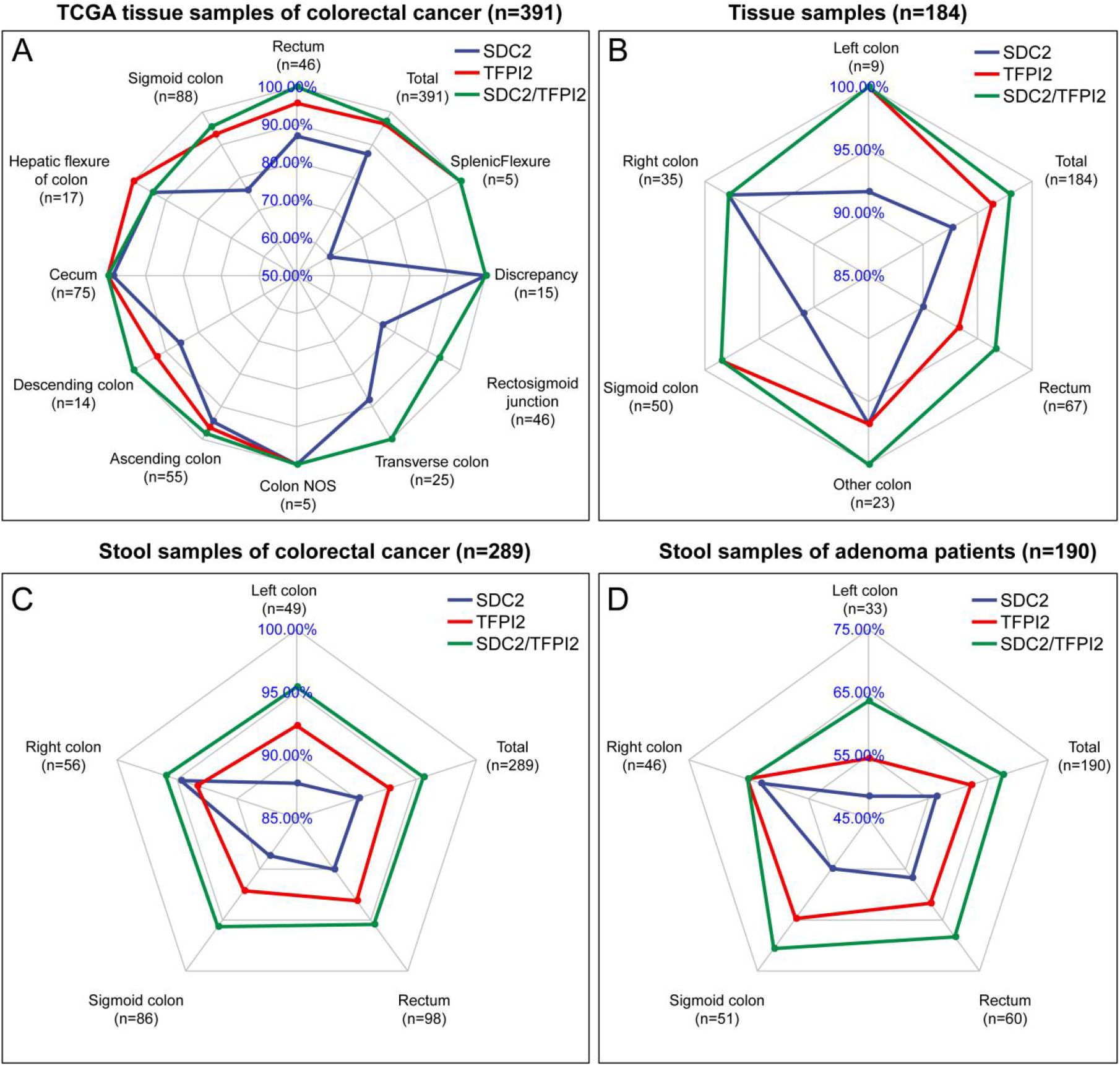
Comparison of the sensitivity of *SDC2, TFPI2* and *SDC2*/*TFPI2* in detecting CRC or adenomas from various locations. Fig 5A shows the results of 391 TCGA cancer tissue specimens. Fig 5(B-D) shows detection sensitivity of the 184 CRC tissue specimens, 289 CRC stool specimens and 190 adenomas specimens collected in this study, respectively. The cutoff value is β=0.2 for Fig 5A and Ct=38 for Fig 5(B-D).

In 184 CRC tissue samples (Fig 5B), when the cut-off values of *SDC2* and *TFPI2* genes were both set to Ct=38, the overall sensitivity of *SDC2*/*TFPI2* was 97.3% while 90.2% for *SDC2*, the difference was highly significant (p≤0.01). Detection rate of cancer from rectum (n=67, *SDC2*/*TFPI2*=95.5%, *SDC2*=86.6%), sigmoid colon (n=50, *SDC2*/*TFPI2*=98.8%, *SDC2*=88.0%) as well as left colon (n=7) showed the most significant improvement.

In 289 CRC stool samples (Fig 5C), when the cut-off values of *SDC2* and *TFPI2* genes were both set to Ct=38, the overall sensitivity of *SDC2*/*TFPI2* was 94.1%, while that of *SDC2* was 86.9%, the difference between *SDC2*/*TFPI2* and *SDC2* was significant (P ≤ 0.001). Left colon (n=49, *SDC2*/*TFPI2*=93.9%, *SDC2*=83.7%), sigmoid colon (n=86, *SDC2*/*TFPI2*=94.2%, *SDC2*=84.9%) and rectum (n=98, *SDC2*/*TFPI2*=93.9%, *SDC2*=86.7%) showed the most significant improvement.

In 190 colorectal adenomas stool samples (Fig 5D) when the cut-off values of *SDC2* and *TFPI2* genes were both set to Ct=38, the sensitivity was 67.4% for *SDC2*/*TFPI2* while 56.3% for *SDC2*, the difference was also highly significant (P≤0.001). Left colon (n=33, *SDC2*/*TFPI2*=66.6%, *SDC2*=48.5%), sigmoid colon (n=51, *SDC2*/*TFPI2*=70.6%, *SDC2*=54.9%) and rectum (n=60, *SDC2*/*TFPI2*=68.3%, *SDC2*=56.7%) showed the most obvious sensitivity improvement.

Based on the above results, it was found that the combined test of *SDC2* and *TFPI2* could reduce the missed rate of CRC and adenomas. 55.3% CRC and 25.3% adenomas specimens with *SDC2* Ct value larger than 38 (false-negative) were *TFPI2* positive (Table 3). *TFPI2* could enhance the sensitivity of *SDC2* especially for cancer in the left colon, rectum and sigmoid colon, and tissue and stool specimens have consistent results.

## 4 Discussion

CRC is an ideal target for intervention since most colorectal tumors grow slowly and remain silent or asymptomatic until they reach a certain size [26]. DNA methylation is an early event and runs through the whole process of tumor development. More and more products use methylated genes as detection targets for CRC, for example, the first blood-based product Epi proColon which targets methylated SEPT9, having a sensitivity of 68%-72% and a specificity of 80% for CRC [27,28]. Recently, Niu et al showed that the detection rate of *SDC2* methylation for CRC and adenomas was 81.1% and 58.2%, and the specificity was 93.3% [21], these sensitivities have room for improvement since the primary aim of screening is to detect neoplasm as much as possible. Our study made a breakthrough in significantly improving the detection rates by adding *TFPI2* to the *SDC2* test system. Although it might have the risk of reducing specificity, the results showed that it still remained at a high level, comparable to that of *SDC2* one target system (Table 2). The sensitivity for CRC and adenomas was 96.6% and 80.0%, while the specificity was 96.40% and 95.7%, respectively. The detection accuracy has been greatly improved compared with Niu et al’s research [21], especially the sensitivities (96.6% vs. 81.1% for CRC, 80.0% vs. 58.2% for adenomas). Of note, both the sensitivities and specificities in our study were higher than that of a Korean study [23], in which the overall sensitivity of CRC, advanced and non-advanced adenomass was 90.2%, 66.7% and 24.4%, the specificity was 90.2%, however, they used two rounds of PCR to amplify methylated *SDC2*, which was relatively cumbersome. As for adenomas detection, a previous literature showed that methylated markers *BMP3, NDRG4, VIM*, and *TFPI2* could detect 68%, 76%, 76%, and 88% advanced adenomass (≥ 1 cm) [29], respectively. However, only 25 adenomas specimens were used in that study and it might be inadequate. It was a pity that the clinical information of adenomas specimens in our study was not very clear, many probably were non-advanced and might mask the true sensitivity for advanced adenomass, which meant that the actual detection rate might be higher than 80.0% in our research.

Analysis of specimens from the TCGA database implied that *SDC2* might have high miss rate for left colon cancer since the methylation level were lower in sigmoid colon cancer, descending colon cancer, and rectosigmoid junction cancer (Table 1). The results of 184 CRC tissue, 289 CRC and 190 adenomas stool specimens also confirmed the conclusions (Fig 5). When *SDC2* was detected in combination with *TFPI2*, the detection rates of cancer increased no matter what the tumor sites were, and the improvement was the most obvious in left colon cancer, sigmoid colon cancer, and rectum cancer. Some samples collected in this study were only recorded that they were from left colon, without specific sources, but it did not have much impact on the conclusion since the sigmoid colon and rectum both belong to the left colon. These results suggest that *SDC2* might have different detection effects on CRC in different parts. Researches showed that the left and right colon had different developmental origins, cancer in these two sites was very different in terms of carcinoma mechanism and prognosis [30]. Sigmoid colon cancer and rectal cancer have a high incidence worldwide, especially in China, they are the two most common CRC types in China [31], the sensitivity improvement in the two biopsy would be of great benefit to the overall CRC detection.

Despite that many methylation-based methods for CRC diagnosis have been reported, there existed some highlights in our study.

- We identified *TFPI2* through whole-genome screening, significantly outperforming the well-established biomarker *SDC2* in CRC detection.
- Five populations from Asian and Euro-America regions and two specimen types (tissue and stool) were involved (Supplemental Table 6), totally covering 1034 patients of CRC, 367 of adenomas, and 190 normal individuals, including different colorectal sites and stages.
- Three indexes (β value, Ct value, ΔCt value) were evaluated (Fig 2, Fig 5 and Table 2), Ct value was a suitable indicator for procedure, being simple to operate and having better performance than ΔCt value.
- *TFPI2* improved the sensitivity of colorectal cancer and adenomas, significantly reduced the proportion of false positive and false negative specimens in *SDC2* assay, whether it is based on the result of the Yoden Index (Table 2) or the result of setting the cut-off value at 38 (Fig 5). Nevertheless, some aspects need further studied in the future:
- Detailed mechanisms of *SDC2*/*TFPI2* hypermethylation in the carcinogenesis should be explored.
- The performance of *SDC2*/*TFPI2* detection in screening and diagnosis of CRC and adenomas should be further evaluated.
- The present assay can be further improved, for example, by involving adenomas-specific markers to help detection of CRC and precancerous lesions.

As the most commonly used non-invasive detection methods in clinical practice at present, FOBT and FIT have relatively limited detection accuracy, the sensitivity for colorectal adenomas is less than 30% [32], our approach provided much better performance. As for other method such as CT colonography, there’s research showing that its precision is comparable to that of optical endoscopy [33], and study showing that it’s inferior to the latter [32], the method established in this study provided results highly consistent with endoscopy/pathology. It is stool-based, which is safe and operates easily, avoiding bowel preparation and possible cross-infection during colonoscopy, also avoiding radiation during CT colonography. Cologuard is a kind of multi-target stool DNA detection kit for CRC screening through the combined detection of KRAS mutations, DNA methylation (NDRG4 and BMP3) and hemoglobin. However, the detection technologies are complicated which involve gene mutation, DNA methylation and protein detection, also the kit is very expensive, limiting its use in the screening of CRC. However, our research only used MSP technology to detect the DNA methylation of *SDC2* and *TFPI2*, so it was simpler and cheaper than Cologuard.

## Conclusion

Our research has proved that *SDC2* and *TFPI2* gene methylation had complementary effects in CRC screening. The combination of the two had high diagnostic value for CRC, with extremely high sensitivity and specificity. Therefore, the combined of *SDC2* and *TFPI2* DNA methylation provided a feasible method for early screening of CRC.

## Data Availability

Please contact the corresponding author for data requests.

## 5 Author Contributions

Lei Yin, Lianglu Zhang, and Jun Lin contributed to the conception and design. Lanlan Dong contributed to the specimen collection, experiment, and data analysis, Changming Lu contributed to the methodology, assembly of data. Wenxian Huang, Cuiping Yang, Qian Wang, Ruixue Lei, Rui Sun contributed to the specimen collection and clinical information collection. Kangkang Wan, Tingting Li, Fan Sun, and Tian Gan contributed to experiment and data analysis. All authors helped to the data interpretation and manuscript writing and reviewing. All authors agreed to the final manuscript.

## Acknowledgments

This research is supported by the Department of Health and Family Planning Commission of Jiading District (Grants 2008-ky-01) and Ruijin North Hospital (Grants 2018 zy16). The study was also supported, in part, by grants from the Natural Science Foundation of China (No. 81502461).

## Reference

1. Bray F, Ferlay J, Soerjomataram I, et al. Global cancer statistics 2018: GLOBOCAN estimates of incidence and mortality worldwide for 36 cancers in 185 countries. CA Cancer J Clin. 2018; 68:394–424.

2. Zheng RS, Sun KX, Zhang SW, et al. Report of cancer epidemiology in China, 2015. Zhonghua Zhong Liu Za Zhi. 2019; 41:19–28.

3. Lan YT, Yang SH, Chang SC, et al. Analysis of the seventh edition of American Joint Committee on colon cancer staging. Int J Colorectal Dis. 2012; 27:657–63.

4. Grady WM, Markowitz SD. Genetic and epigenetic alterations in colon cancer. Annual review of genomics and human genetics. 2002; 3:101–28.

5. Chung DC. Genetic Testing and Early Onset Colon Cancer. Gastroenterology. 2018; 154:788–9.

6. Kahi CJ, Imperiale TF, Juliar BE, et al. Effect of screening colonoscopy on colorectal cancer incidence and mortality. Clin Gastroenterol Hepatol. 2009; 7:770–5.

7. Vermeer NC, Snijders HS, Holman FA, et al. Colorectal cancer screening: Systematic review of screen-related morbidity and mortality. Cancer Treat Rev. 2017; 54:87–98.

8. Werner S, Krause F, Rolny V, et al. Evaluation of a 5-Marker Blood Test for Colorectal Cancer Early Detection in a Colorectal Cancer Screening Setting. Clin Cancer Res. 2016; 22:1725–33.

9. Grady WM, Pritchard CC. Molecular alterations and biomarkers in colorectal cancer. Toxicol Pathol. 2014; 42:124–39.

10. Yiu AJ, Yiu CY. Biomarkers in Colorectal Cancer. Anticancer Res. 2016; 36:1093–102.

11. Novak P, Jensen TJ, Garbe JC, et al. Stepwise DNA methylation changes are linked to escape from defined proliferation barriers and mammary epithelial cell immortalization. Cancer Res. 2009; 69:5251–8.

12. Müller HM, Oberwalder M, Fiegl H, et al. Methylation changes in faecal DNA: a marker for colorectal cancer screening? Lancet. 2004; 363:1283–5.

13. Loh K, Chia JA, Greco S, et al. Bone morphogenic protein 3 inactivation is an early and frequent event in colorectal cancer development. Genes Chromosomes Cancer. 2008;47: 449–60.

14. deVos T, Tetzner R, Model F, et al. Circulating methylated SEPT9 DNA in plasma is a biomarker for colorectal cancer. Clin Chem. 2009; 55:1337–46.

15. Melotte V, Lentjes MH, van den Bosch SM, et al. N-Myc downstream-regulated gene 4 (NDRG4): a candidate tumor suppressor gene and potential biomarker for colorectal cancer. J Natl Cancer Inst. 2009; 101:916–27.

16. Oh T, Kim N, Moon Y, et al. Genome-wide identification and validation of a novel methylation biomarker, SDC2, for blood-based detection of colorectal cancer. J Mol Diagn. 2013;15: 498-507.

17. Robertson DJ, Imperiale TF. Stool Testing for Colorectal Cancer Screening. Gastroenterology. 2015; 149:1286–93.

18. Mojtabanezhad Shariatpanahi A, Yassi M, Nouraie M, et al. The importance of stool DNA methylation in colorectal cancer diagnosis: A meta-analysis. PLoS One. 2018; 13:e0200735.

19. Imperiale TF, Ransohoff DF, Itzkowitz SH, et al. Multitarget stool DNA testing for colorectal-cancer screening. N Engl J Med. 2014; 370:1287–97.

20. Mitchell SM, Ross JP, Drew HR, et al. A panel of genes methylated with high frequency in colorectal cancer. BMC Cancer. 2014; 14:54.

21. Niu F, Wen J, Fu X, et al. Stool DNA Test of Methylated Syndecan-2 for the Early Detection of Colorectal Neoplasia. Cancer Epidemiol Biomarkers Prev. 2017; 26:1411–9.

22. Oh TJ, Oh HI, Seo YY, et al. Feasibility of quantifying SDC2 methylation in stool DNA for early detection of colorectal cancer. Clin Epigenetics. 2017; 9:126.

23. Han YD, Oh TJ, Chung TH, et al. Early detection of colorectal cancer based on presence of methylated syndecan-2 (SDC2) in stool DNA. Clin Epigenetics. 2019; 11:51.

24. Luo Y, Wong CJ, Kaz AM, et al. Differences in DNA methylation signatures reveal multiple pathways of progression from adenoma to colorectal cancer. Gastroenterology. 2014; 147:418–29.

25. Alvi MA, Loughrey MB, Dunne P, et al. Molecular profiling of signet ring cell colorectal cancer provides a strong rationale for genomic targeted and immune checkpoint inhibitor therapies. Br J Cancer. 2017; 117:203–9.

26. Vogelstein B, Fearon ER, Hamilton SR, et al. Genetic alterations during colorectal-tumor development. N Engl J Med. 1988; 319:525–32.

27. Johnson DA, Barclay RL, Mergener K, et al. Plasma Septin9 versus fecal immunochemical testing for colorectal cancer screening: a prospective multicenter study. PLoS One. 2014; 9:e98238.

28. Nian J, Sun X, Ming S, et al. Diagnostic Accuracy of Methylated SEPT9 for Blood-based Colorectal Cancer Detection: A Systematic Review and Meta-Analysis. Clinical and translational gastroenterology. 2017; 8:e216.

29. Zou H, Allawi H, Cao X, et al. Quantification of methylated markers with a multiplex methylation-specific technology. Clin Chem. 2012; 58:375–83.

30. Meza R, Jeon J, Renehan AG, et al. Colorectal cancer incidence trends in the United States and United kingdom: evidence of right-to left-sided biological gradients with implications for screening. Cancer Res. 2010; 70:5419–29.

31. Li M, Gu J. Changing patterns of colorectal cancer in China over a period of 20 years. World J Gastroenterol. 2005; 11:4685–8.

32. Graser A,Stieber P, Nagel D, et al. Comparison of CT colonography, colonoscopy, sigmoidoscopy and faecal occult blood tests for the detection of advanced adenoma in an average risk population. Gut. 2009; 58:241–8.

33. Kim DH, Pickhardt PJ, Taylor AJ, et al. CT colonography versus colonoscopy for the detection of advanced neoplasia. N Engl J Med. 2007; 357:1403–12.

