## Supplemental 1-6 for "Diagnostic value in colorectal tumorous lesions of *SDC2/TFPI2* methylation based on bowel subsite difference"

**Supplemental Table 1. Clinicopathological features of colorectal cancerous and normal tissues**

| Characteristics | Cancer | Control |
| --- | --- | --- |
|  | (n=184) | (n=54) |
| Sex, no.(%) |  |  |
| Male | 112 (60.9%) | 32 (59.3%) |
| Female | 72 (39.1%) | 22 (40.7%) |
| Age, Median (range) | 63(30-94) | 59 (32-87) |
| Localization, no. (%) |  |  |
| Colon |  |  |
| Left colon | 9 (4.9%) | 1 (1.9%) |
| Right colon | 35 (19.0%) | 15 (27.8%) |
| Sigmoid colon | 50 (27.2%) | 24 (44.4%) |
| Other colon | 23 (12.5%) | - |
| Rectum | 67 (36.4%) | 14 (25.9%) |
| Differentiation grade, no.(%) |  |  |
| Low | 21 (11.4%) | - |
| Moderate | 137 (74.5%) | - |
| High | 14 (7.6%) | - |
| NA | 12 (6.5%) | - |

**Supplemental Table 2. Clinicopathological features of colorectal cancer, adenoma and normal stool specimens**

| Characteristics | Carcinoma | Adenoma | Normal |
| --- | --- | --- | --- |
|  | (n=289) | (n=190) | (n=217) |
| Sex, no.(%) |  |  |  |
| Male | 156 (54.0%) | 112 (58.9%) | 131 (60.4%) |
| Female | 133 (46.0%) | 78 (41.1%) | 86 (39.6%) |
| Age, Median (range) | 61 (22-81) | 61 (22-89) | 59 (29-81) |
| Localization, no. (%) |  |  |  |
| Left colon | 49 (17.0%) | 33 (17.4%) | - |
| Right colon | 56 (19.4%) | 46 (24.2%) | - |
| Sigmoid colon | 86 (29.8%) | 51 (26.8%) | - |
| Rectal | 98 (33.8%) | 60 (31.6%) | - |

**Supplemental Table 3. Sequences of primers and probes used in this study**

| Target | Description | Sequence (5’-3’) |
| --- | --- | --- |
| *ACTB* | Capture probe | TGGGTCTGCGCTGTAAGAGTTGGTTGCC |
|  | MSP Forward Primer | AAGGTGGTTGGGTGGTTGTTTTG |
|  | MSP Reverse Primer | AATAACACCCCCACCCTGC |
|  | MSP Probe | GGAGTGGTTTTTGGGTTTG |
| *SDC2* | Capture probe | AATCCGGAGCAGAGTACCGCAGCGA TT |
|  | MSP Forward Primer | CGAGTTTGAGTCGTAATCGTTGC |
|  | MSP Reverse Primer | TCCGCCGACACGCAAACCACCAAA CC |
|  | MSP Probe | AACAAAACGAAACCTCCTACCCAA C |
| *TFPI2* | Capture probe | CTGTCGTAGTAGTAACGGAGAAGTAGGG C |
|  | MSP Forward Primer | CGCGGAGATTTGTTTTTTGT |
|  | MSP Reverse Primer | AACAAACATCGTCGCAAACCT C |
|  | MSP Probe | ATAAAACCCGACAAAATCCG |

**Supplemental Table 4. Demographic characteristics of patients retrieved from TCGA database**

| Patient characteristics | Cancer group (n=391) | Control group (n=45) |
| --- | --- | --- |
| Sex, no. (%) |  |  |
| Male | 211 (54.0%) | 24 (53.3%) |
| Female | 180 (46.0%) | 21 (46.7%) |
| Age, Median (range) | 66 (31-90) | 62 (34-84) |
| Race, no. (%) |  |  |
| White | 282 (72.1%) | 20 (44.4%) |
| Black /African American | 63 (16.1%) | 4 (8.9%) |
| American Indian/Alaska native | 1 (0.3%) | - |
| Asian | 12 (3.1%) | - |
| Not Reported | 33 (8.4%) | 21 (46.7%) |
| Localization, no. (%) |  |  |
| Rectum NOS | 49 (12.5%) | 6 (13.3%) |
| Sigmoid colon | 67 (17.1%) | 7 (15.6%) |
| Hepatic flexure of colon | 13 (3.3%) | 3 (6.7%) |
| Cecum | 71 (18.2%) | 7 (15.6%) |
| Descending colon | 14 (3.6%) | 2 (4.4%) |
| Ascending colon | 60 (15.3%) | 6 (13.3%) |
| Colon NOS | 49 (12.5%) | 12 (26.7%) |
| Transverse colon | 14 (3.6%) | - |
| Rectosigmoid junction | 46 (11.8%) | 2 (4.4%) |
| Others | 8 (2.1%) | - |
| UICC* stage, no. (%) |  |  |
| I | 55 (14.1%) | - |
| II | 143 (36.6%) | - |
| III | 120 (30.7%) | - |
| IV | 54 (13.8%) | - |
| Not Reported | 19 (4.9%) | - |

*UICC means Union of International Cancer Control

**Supplemental Table 5. Colorectal cancer specimens with SDC2 β-value lower than 0.2 selected from TCGA database**

| No. | Specimen ID | Tumor location | SDC2 | TFPI2 |
| --- | --- | --- | --- | --- |
| 1 | TCGA-DM-A1D6-01A-21D-A153-05 | Splenic flexure of colon | 0.050 | 0.784 |
| 2 | TCGA-DM-A1D0-01A-11D-A153-05 | Sigmoid colon | 0.060 | 0.506 |
| 3 | TCGA-DY-A1DC-01A-31D-A153-05 | Rectosigmoid junction | 0.060 | 0.617 |
| 4 | TCGA-A6-6142-01A-11D-1772-05 | Sigmoid colon | 0.062 | 0.164 |
| 5 | TCGA-A6-2675-01A-02D-1721-05 | Sigmoid colon | 0.064 | 0.293 |
| 6 | TCGA-G4-6307-01A-11D-1721-05 | Sigmoid colon | 0.065 | 0.558 |
| 7 | TCGA-A6-6140-01A-11D-1772-05 | Descending colon | 0.065 | 0.739 |
| 8 | TCGA-F5-6863-01A-11D-1926-05 | Rectum, NOS | 0.066 | 0.382 |
| 9 | TCGA-G5-6572-01A-11D-1828-05 | Rectosigmoid junction | 0.067 | 0.650 |
| 10 | TCGA-CM-6164-01A-11D-1651-05 | Sigmoid colon | 0.067 | 0.538 |
| 11 | TCGA-AZ-6603-01A-11D-1837-05 | Sigmoid colon | 0.069 | 0.575 |
| 12 | TCGA-CA-6715-01A-21D-1837-05 | Sigmoid colon | 0.070 | 0.759 |
| 13 | TCGA-A6-2685-01A-01D-1407-05 | Colon, NOS | 0.071 | 0.332 |
| 14 | TCGA-CM-6162-01A-11D-1651-05 | Ascending colon | 0.073 | 0.154 |
| 15 | TCGA-CA-5797-01A-01D-1651-05 | Colon, NOS | 0.073 | 0.195 |
| 16 | TCGA-BM-6198-01A-11D-1734-05 | Rectum, NOS | 0.078 | 0.496 |
| 17 | TCGA-D5-5541-01A-01D-1651-05 | Sigmoid colon | 0.084 | 0.683 |
| 18 | TCGA-DC-6157-01A-11D-1658-05 | Rectosigmoid junction | 0.096 | 0.330 |
| 19 | TCGA-AF-6672-01A-11D-1828-05 | Rectosigmoid junction | 0.098 | 0.277 |
| 20 | TCGA-CM-6674-01A-11D-1837-05 | Hepatic flexure of colon | 0.098 | 0.375 |
| 21 | TCGA-5M-AATE-01A-11D-A40X-05 | Ascending colon | 0.102 | 0.666 |
| 22 | TCGA-A6-3810-01A-01D-A27A-05 | Colon, NOS | 0.106 | 0.545 |
| 23 | TCGA-DY-A0XA-01A-11D-A153-05 | Rectosigmoid junction | 0.107 | 0.318 |
| 24 | TCGA-AA-3494-01A-01D-1407-05 | Colon, NOS | 0.112 | 0.258 |
| 25 | TCGA-A6-A567-01A-31D-A28O-05 | Sigmoid colon | 0.119 | 0.483 |
| 26 | TCGA-F5-6810-01A-11D-1828-05 | unknown | 0.122 | 0.268 |
| 27 | TCGA-A6-A565-01A-31D-A28O-05 | Transverse colon | 0.122 | 0.234 |
| 28 | TCGA-DC-6158-01A-11D-1658-05 | Rectum, NOS | 0.122 | 0.442 |
| 29 | TCGA-DM-A1DB-01A-11D-A153-05 | Sigmoid colon | 0.125 | 0.374 |
| 30 | TCGA-DC-4749-01A-01D-1734-05 | Rectosigmoid junction | 0.134 | 0.382 |
| 31 | TCGA-CL-5917-01A-11D-1658-05 | Rectosigmoid junction | 0.135 | 0.503 |
| 32 | TCGA-EI-6509-01A-11D-1734-05 | Rectum, NOS | 0.139 | 0.305 |
| 33 | TCGA-F5-6465-01A-11D-1734-05 | Rectosigmoid junction | 0.140 | 0.312 |
| 34 | TCGA-DM-A28E-01A-11D-A16X-05 | Sigmoid colon | 0.147 | 0.293 |
| 35 | TCGA-A6-5662-01A-01D-1651-05 | Splenic flexure of colon | 0.148 | 0.650 |
| 36 | TCGA-AY-A8YK-01A-11D-A40X-05 | Sigmoid colon | 0.149 | 0.285 |
| 37 | TCGA-AD-6964-01A-11D-1926-05 | Ascending colon | 0.151 | 0.339 |
| 38 | TCGA-AZ-5403-01A-01D-1651-05 | Descending colon | 0.156 | 0.285 |
| 39 | TCGA-F4-6703-01A-11D-1837-05 | Ascending colon | 0.156 | 0.109 |
| 40 | TCGA-DM-A1D7-01A-11D-A153-05 | Sigmoid colon | 0.162 | 0.410 |
| 41 | TCGA-5M-AAT6-01A-11D-A40X-05 | Ascending colon | 0.166 | 0.251 |
| 42 | TCGA-AF-A56K-01A-32D-A39G-05 | Rectosigmoid junction | 0.180 | 0.160 |
| 43 | TCGA-AF-6655-01A-11D-1828-05 | Rectosigmoid junction | 0.180 | 0.140 |
| 44 | TCGA-D5-6923-01A-11D-1926-05 | Sigmoid colon | 0.181 | 0.512 |
| 45 | TCGA-D5-6932-01A-11D-1926-05 | Colon, NOS | 0.182 | 0.585 |
| 46 | TCGA-AZ-4308-01A-01D-1407-05 | Sigmoid colon | 0.189 | 0.470 |
| 47 | TCGA-CI-6621-01A-11D-1828-05 | Rectum, NOS | 0.191 | 0.481 |
| 48 | TCGA-AH-6643-01A-11D-1828-05 | Rectosigmoid junction | 0.191 | 0.753 |
| 49 | TCGA-CM-6170-01A-11D-1651-05 | Descending colon | 0.198 | 0.619 |
| 50 | TCGA-F5-6812-01A-11D-1828-05 | Connective, subcutaneous and other soft tissues of abdomen | 0.136 | 0.283 |

**Supplemental Table 6. Description of the specimen groups used in this study**

| Specimen group | TCGA | GSE48684 | GSE79740 | D184 | D289 |
| --- | --- | --- | --- | --- | --- |
| Specimen type | Tissue | Tissue | Tissue | Tissue | Stool |
| Data source | GPL13534 | GPL13534 | GPL13534 | Collected in this study | Collected in this study |
| Method^a,b^ | 450k | 450k | 450k | PCR | PCR |
| Methylation indicator | β values | β values | β values | Ct values | Ct values |
| Normal specimens | 45 | 41 | 10 | 54 | 217 |
| Adenoma specimens | 0 | 0 | 0 | 0 | 190 |
| CRC specimens | 411 | 106 | 44 | 184 | 289 |
| Demographic feature^c^ | Table S4 | NA | NA | Table S1 | Table S2 |
| Use in this study for | Marker discovery | Marker validation | Marker validation | Clinical evaluation | Clinical evaluation |

a.450k means HumanMethylation450 BeadChip.

b. PCR here means methylation-specific PCR developed in this study

c.NA means not available here.
